# Mental health problems, stressful life events and relapse in urinary incontinence in primary school-age childhood: A prospective cohort study

**DOI:** 10.1101/2022.08.12.22278713

**Authors:** Naomi Warne, Jon Heron, Alexander von Gontard, Carol Joinson

## Abstract

Emotional/behaviour problems and exposure to stressful life events are thought to contribute to relapses in urinary incontinence (UI) amongst children who have attained bladder control. However, very few prospective studies have examined these associations. We assessed whether mental health problems and stressful life events were associated with subsequent relapse in UI using multivariable logistic regression in a prospective UK cohort (n=6,408). Mothers provided information on their child’s symptoms of common mental disorders (Development and Wellbeing Assessment, 7 years), stressful life events (7-8 years) and wetting (day and night, 9 years). There was strong evidence that separation anxiety symptoms were associated with UI relapse in the fully adjusted model (OR (95% CI) = 2.08 (1.39, 3.13), p<0.001). Social anxiety, attention deficit hyperactivity disorder and oppositional defiant disorder symptoms were associated with UI relapse, but these associations attenuated following adjustment for child developmental level and earlier emotional/behaviour problems. There was weak evidence for a sex interaction with stressful life events (p=0.065), such that females experiencing more stressful life events were at higher risk of UI relapse (fully adjusted model OR (95% CI) = 1.66 (1.05, 2.61), p=0.029) but there was no association in males (fully adjusted model OR (95% CI) = 0.87 (0.52, 1.47), p=0.608). These results suggest that early treatment of separation anxiety and intervening to reduce the negative outcomes associated with stressful life events (in girls) may help to reduce risk of UI relapse.

## Introduction

Urinary incontinence (UI) is a common problem in primary school-aged children, with bedwetting (enuresis) affecting around 15% and daytime wetting affecting around 8% of 7-year-olds [1, 2]. Most cases of paediatric UI arise from functional impairments in the bladder and are not due to structural, anatomical or neurological causes [3]. Comorbidity of paediatric UI with emotional/behaviour problems is well-established [4]. There is also strong evidence for prospective associations between emotional/behaviour problems and stressful life events in early childhood and problems attaining bladder control at school-age [5, 6]. It is often suggested in resources aimed at clinicians and parents that emotional/behaviour problems and exposure to stressful life events might also contribute to relapses in UI amongst children who have attained bladder control, but there has been little empirical research directed at examining this. *Secondary enuresis* refers to relapses in bedwetting in children who have previously been dry for at least 6 months, whilst *primary enuresis* refers to children who have never achieved 6 months of bladder control [7]. A birth cohort study found that by age 10, almost 8% of children who had previously acquired bladder control had experienced secondary enuresis [8]. Relapses in daytime wetting are also common in primary school-age children [9], although primary and secondary daytime wetting are not differentiated in current classification systems [10]. Cross-sectional studies with small sample sizes have found evidence that children with secondary enuresis experience more stressful life-events when compared to controls without enuresis [9] and children with primary enuresis [11]. Also, children with secondary enuresis have more comorbid emotional/behavioural problems than children with primary enuresis [12].

Birth cohort studies conducted in New Zealand in the 1980s and 1990s provide evidence for cross-sectional and prospective associations between emotional/behaviour problems, stressful life events and secondary enuresis. McGee et al. (1984) found higher parent-reported behaviour problem scores at ages 7 and 9 in 9-year-old children with secondary enuresis (defined by the authors as involuntary voiding of urine by day or night at least once a month) compared with children without enuresis [13]. Feehan et al. (1990) found higher parent-reported behaviour problem scores and self-reported anxiety scores in 11-year-old children with secondary enuresis (defined as day or night wetting at least once a month with a dry period of at least 1 year prior to onset) compared with dry children at age 11, as well as at the age of 13 years [14]. This implies that secondary enuresis is associated with long-term effects on mental health. Fergusson et al. (1990) found that the risk of secondary enuresis at age 10 (defined as onset of bedwetting after age 5 following at least 1 year of remaining dry at night) increased with the child’s level of exposure to adverse family life events during the period from 4 to 10 years [8]. The study also found that children who were exposed to four or more life events in a given year had an increased risk of secondary enuresis in that year compared with unexposed children. Jarvelin et al. (1990) found secondary enuretic children had significantly more weighted life events at the age of 6 years [15]. There was a significant accumulation of life events in the year before the onset of secondary wetting, but not over the 2 years before the relapse occurred. The authors of these previous birth cohort studies argue that emotional/behavioural problems and stressful life events might precipitate a relapse in wetting, but there are limitations that should be considered when interpreting the findings. One study examined cross-sectional associations between emotional/behaviour problems and secondary enuresis at age 11, leading to the possibility of reverse causality as an alternative explanation for their findings [14]. Only one of the studies adjusted for confounders (child’s sex, perinatal history and family socioeconomic background) but they did not adjust for the child’s developmental level, nor did they examine whether emotional/behaviour problems in early childhood are associated with secondary enuresis [8]. Loss to follow up was also a limitation of this study, such that families of lower socioeconomic background were underrepresented in the study sample leading to potential selection bias.

The present study, based on data from a large UK birth cohort, examined whether symptoms of common mental health disorders in 7-year-old children who were dry during the day and night were associated with an increased risk of relapse in UI (bedwetting and/or daytime wetting) at age 9. The study also examined whether stressful life events occurring when children were 7-8 years-old were associated with an increased risk of relapse in UI at age 9.

## Methods

### Participants

We used data from the Avon Longitudinal Study of Parents and Children (ALSPAC) [16, 17]. This cohort recruited pregnant women in the Avon area of Bristol, UK, with an expected delivery date between 1^st^ April 1991 and 31^st^ December 1992 (14,541 pregnancies enrolled with at least one returned questionnaire or attended face-to-face clinic by 19^th^ July 1999) and has followed the families to present day. Of the 14,676 foetuses, there were 14,062 live births and 13,988 were alive at 1 year. Please note that the study website contains details of all the data that are available through a fully searchable data dictionary and variable search tool: http://www.bristol.ac.uk/alspac/researchers/our-data/. We used an imputed sample of 6,408 children (3,143 males, 3,265 females) from the core sample who were dry during the day and night at 7 years (Figure 1). Dry at 7 years was defined as maternal responses of “never” to both “How often does your child wet him/herself during the day?” and “How often does your child wet the bed at night?”. Differences between those who were included in this sample and excluded (wet at 7 years) are presented in Table S1.

**Fig 1.**
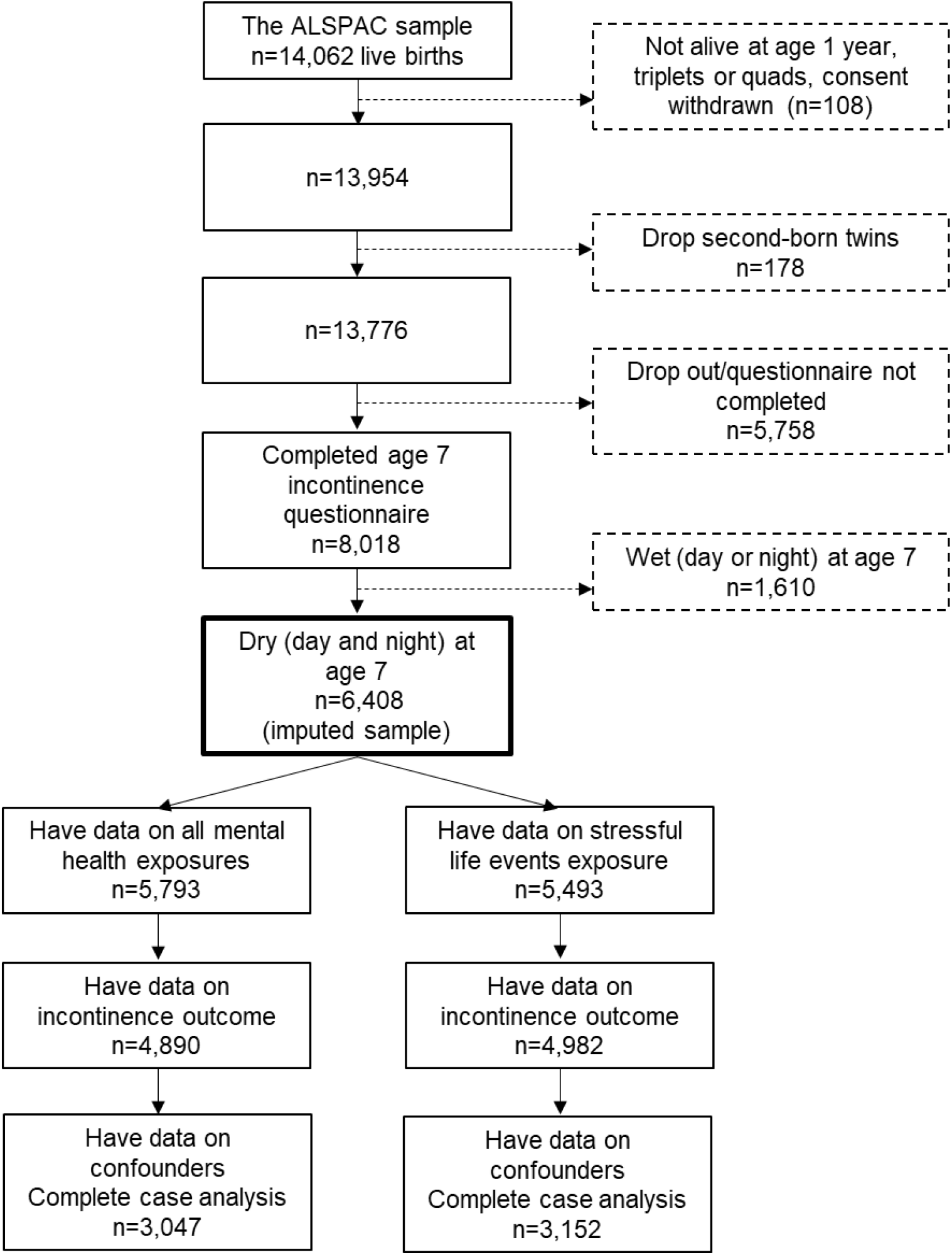
ALSPAC flowchart of attrition and sample derivation Note: ALSPAC – Avon Longitudinal Study of Parents and Children

### Ethical approval

Ethical approval for the study was obtained from the ALSPAC Ethics and Law Committee and the Local Research Ethics Committees. Informed consent for the use of data collected via questionnaires and clinics was obtained from participants following the recommendations of the ALSPAC Ethics and Law Committee at the time.

### Exposures

Symptoms of common mental health disorders were measured at 7 years based on maternal reports on the Development and Wellbeing Assessment (DAWBA) [18]. We included symptoms of separation anxiety, phobias, social anxiety, post-traumatic stress disorder (PTSD), obsessive compulsive disorder (OCD), generalised anxiety disorder (GAD), depression, attention deficit hyperactivity disorder (ADHD), oppositional defiant disorder (ODD) and conduct disorder. A small number of children met DSM-IV criteria for psychiatric disorders in the ALSPAC cohort at this age [19, 20]. We, therefore, created binary variables to indicate the presence of any symptom that was severe, e.g. rated as “a lot more than others” or “a great deal” (Table S2).

Main carer (typically maternal) reports of child-based stressful life events occurring when the child was 7-8 years old were obtained from a questionnaire completed when the study child was 8.5 years. Mothers were asked whether a range of life events had occurred and the impact since the child’s 7^th^ birthday including: being taken into care; a pet dying; moving home; being physically hurt by someone; being sexually abused; having a family member die; separation from mother, father or someone else; having a new mother, father or sibling; being admitted to hospital; having a change in caretaker; starting a new school; and losing a best friend. Summed life events were dichotomised into a binary score of three or more events (1) versus two or fewer events (0).

### Outcome

Mothers reported on two questions assessing the frequency that their child wet him/herself during the day and wet the bed at night at 9 years (identical questions to those asked at age 7). A binary variable was derived which defined the presence of any wetting during the day and/or night as a relapse in wetting (responses of “occasional accident but less than once a week”, “about once a week”, “2-5 times a week”, “nearly every day”, and “more than once a day”).

### Confounders

We adjusted for a range of confounders sequentially in analyses including (i) sex (0 male, 1 female); (ii) indicators of socioeconomic disadvantage measured around birth (low parental social class, low maternal age at birth, higher parity, lower maternal education) and close to exposure measurement (insecure housing tenure and material hardship at 5 years, major financial difficulties and lack of social support at 6 years); (iii) poor maternal mental health (maternal depression and anxiety symptoms) at 6 years; and (iv) child development at 18 months and emotional/behaviour problems at 6.75 years. Analyses with mental health symptoms were also adjusted for early child stressful life events (5-6 years) in a subsequent step. Full information on confounders and their derivation is available in Table S3.

### Statistical Analysis

All analyses were performed in Stata version 16 [21]. First, we conducted descriptive statistics on the exposure and outcome data and examined sex differences in prevalence. We examined the association of mental health symptoms and stressful life event exposures with UI relapse in a series of logistic regressions. We first performed unadjusted analyses, and then examined evidence for sex interactions. Where there was evidence of a sex interaction, we provide results for males and females separately from sex interaction models. Where there was no evidence of a sex interaction, we did not include the sex interaction term in subsequent models. We adjusted for confounders sequentially to allow us to examine the level of attenuation in the effect estimate that was associated with each confounder.

### Missing data

Analyses were conducted on an imputed dataset of 6,408 individuals with complete data on incontinence, and who were dry during the day and night, at 7 years old. We used multivariate imputation by chained equations to impute missing data on exposure, outcome and confounders using the mi impute chained command in Stata [22] under the Missing at Random (MAR) assumption. In addition to variables in the main analysis, we included variables auxiliary variables that were likely related to the missing data mechanism in the imputation model. These included earlier emotional/behavioural problems (4 and 6 years); earlier urinary and faecal incontinence during the day and night (5 and 6 years); presence of urinary tract infection, constipation and urinary symptoms at age 7; earlier child developmental level (6 months); and earlier summed stressful life events score (4-5 years). Earlier measures of key sociodemographic indicators (housing tenure, major financial difficulties, material hardship, social support) and maternal mental health (depression and anxiety) were also included. Two hundred and fifty datasets were imputed (a decision informed by studying the Monte Carlo errors for the estimated parameters). We used a sex-stratified approach to imputation using the by() option.

## Results

### Descriptive results

Descriptive statistics for the exposure and outcome variables are available in Table 1. Prevalence of mental health symptoms ranged from 2.99% (PTSD) to 13.13% (ADHD). Females were more likely to have a phobia symptom or a depression symptom, but males were more likely to have social anxiety, OCD, ADHD, ODD and conduct symptoms. 17.77% of the sample experienced three or more stressful life events and 4.53% had daytime or night-time wetting at 9 years. Comparison between observed and imputed samples for these variables is available in Table S4.

**Table 1.** Descriptive information on mental health symptoms, stressful life events and wetting in the imputed sample

|  | Total (n=6408) |  | Males (n=3143) |  | Females (n=3265) |  | Sex differences |  |
| --- | --- | --- | --- | --- | --- | --- | --- | --- |
|  | % | SE | % | SE | % | SE | OR (95% CI) | p |
| **Age 7** |  |  |  |  |  |  |  |  |
| *Separation anxiety* | 6.59 | 0.31 | 6.84 | 0.45 | 6.34 | 0.43 | 0.93 (0.77, 1.12) | .417 |
| *Phobia* | 11.56 | 0.40 | 10.45 | 0.55 | 12.62 | 0.58 | 1.21 (1.05, 1.38) | .007 |
| *Social anxiety* | 5.13 | 0.28 | 6.05 | 0.43 | 4.25 | 0.36 | 0.70 (0.57, 0.87) | .001 |
| *PTSD* | 2.99 | 0.21 | 2.77 | 0.29 | 3.20 | 0.31 | 1.16 (0.87, 1.53) | .310 |
| *OCD* | 7.12 | 0.33 | 7.90 | 0.49 | 6.36 | 0.43 | 0.80 (0.67, 0.96) | .018 |
| *GAD* | 8.03 | 0.34 | 8.32 | 0.50 | 7.74 | 0.47 | 0.93 (0.79, 1.10) | .394 |
| *Depression* | 11.12 | 0.40 | 10.21 | 0.55 | 12.00 | 0.58 | 1.17 (1.02, 1.35) | .026 |
| *ADHD* | 13.13 | 0.42 | 16.54 | 0.67 | 9.86 | 0.53 | 0.60 (0.52, 0.68) | <.001 |
| *ODD* | 5.24 | 0.28 | 6.89 | 0.45 | 3.66 | 0.33 | 0.53 (0.43, 0.66) | <.001 |
| *Conduct disorder* | 5.85 | 0.30 | 6.49 | 0.44 | 5.22 | 0.39 | 0.80 (0.66, 0.98) | .031 |
| **Age 7-8** |  |  |  |  |  |  |  |  |
| *Stressful life events* | 17.77 | 0.52 | 17.12 | 0.74 | 18.40 | 0.73 | 1.08 (0.96, 1.21) | .216 |
| **Age 9** |  |  |  |  |  |  |  |  |
| *Wetting (day or night)* | 4.53 | 0.29 | 4.92 | 0.43 | 4.16 | 0.38 | 0.84 (0.66, 1.09) | .187 |

PTSD = Post-Traumatic Stress Disorder; OCD = Obsessive Compulsive Disorder; GAD = Generalised Anxiety Disorder; ADHD = Attention Deficit Hyperactivity
Disorder; ODD = Oppositional Defiant Disorder; SE = standard error

In males who relapsed at age 9 (observed data), 75% were wetting during the night and 22.6% were wetting during the day. In females, 39.8% relapsed with bedwetting and 55.6% relapsed with daytime wetting. We are unable to report combined day and night wetting due to ALSPAC stipulations on small cell counts of 5 or fewer individuals to preserve anonymity.

Amongst children who were dry in the day at age 7 (observed data), 95.3% were dry (4.7% wet) during the day at age 5 and 94.7% were dry (5.3% wet) during the day at age 6. Amongst children who were dry at night at age 7 (observed data), 87.6% were dry (12.4% wet) at age 5 and 91.3% were dry (8.7% wet) at age 6.

### Associations between mental health symptoms at 7 years and wetting at 9 years

Table 2 shows the logistic regression results in the imputed sample. In unadjusted analyses, presence of separation anxiety, social anxiety, ADHD and ODD symptoms were associated with wetting at 9 years (Table 2). There was no evidence of sex interactions (ORs 0.73-1.53, p values > 0.27). There was strong evidence for an association with separation anxiety throughout adjustments for confounders, including in the fully adjusted model (OR (95% CI) = 2.08 (1.39, 3.13), p<0.001). However, the associations with the other symptoms of common mental disorders were attenuated in the fully adjusted models. There was a large attenuation of these effects following adjustment for child developmental level and emotional/behaviour problems in early childhood (results including each step of adjustment are available in Table S5). Results from complete case analyses (Table S6) were largely consistent with imputed results.

**Table 2.** Associations between mental health symptoms at 7 years and relapse of wetting at 9 years (imputed data, n=6408)

| Exposure | Unadjusted |  | Adjusted for sex and socioeconomic disadvantage |  | Adjusted for sex, socioeconomic disadvantage, and maternal mental health |  | Adjusted for sex, socioeconomic disadvantage, maternal mental health, child developmental level, child mental health and child stressful life events |  |
| --- | --- | --- | --- | --- | --- | --- | --- | --- |
|  | OR (95%CI) | p | OR (95%CI) | p | OR (95%CI) | p | OR (95%CI) | p |
| **Separation anxiety** | 2.63 (1.79, 3.85) | <.001 | 2.51 (1.70, 3.71) | <.001 | 2.31 (1.55, 3.44) | <.001 | 2.08 (1.39, 3.13) | <.001 |
| **Phobia** | 1.13 (0.77, 1.67) | .537 | 1.07 (0.72, 1.58) | .750 | 1.02 (0.68, 1.51) | .936 | 0.90 (0.60, 1.35) | .617 |
| **Social anxiety** | 1.96 (1.24, 3.11) | .004 | 1.83 (1.15, 2.92) | .011 | 1.69 (1.06, 2.72) | .029 | 1.47 (0.90, 2.38) | .121 |
| **PTSD** | 1.19 (0.56, 2.52) | .649 | 1.01 (0.47, 2.18) | .976 | 0.91 (0.42, 1.98) | .819 | 0.77 (0.35, 1.70) | .516 |
| **OCD** | 0.94 (0.56, 1.56) | .797 | 0.85 (0.51, 1.42) | .527 | 0.80 (0.48, 1.34) | .393 | 0.66 (0.39, 1.13) | .132 |
| **GAD** | 1.22 (0.78, 1.91) | .377 | 1.16 (0.74, 1.81) | .528 | 1.06 (0.67, 1.67) | .799 | 0.92 (0.58, 1.47) | .731 |
| **Depression** | 1.41 (0.98, 2.05) | .066 | 1.32 (0.90, 1.92) | .151 | 1.23 (0.84, 1.81) | .279 | 1.12 (0.76, 1.65) | .580 |
| **ADHD** | 1.65 (1.18, 2.30) | .003 | 1.49 (1.06, 2.10) | .022 | 1.40 (0.99, 1.98) | .061 | 1.05 (0.71, 1.54) | .822 |
| **ODD** | 2.21 (1.43, 3.42) | <.001 | 1.94 (1.24, 3.04) | .004 | 1.82 (1.15, 2.86) | .010 | 1.35 (0.82, 2.21) | .237 |
| **Conduct disorder** | 1.26 (0.75, 2.13) | .376 | 1.11 (0.65, 1.89) | .702 | 1.03 (0.60, 1.76) | .924 | 0.81 (0.47, 1.41) | .454 |

PTSD = Post-Traumatic Stress Disorder; OCD = Obsessive Compulsive Disorder; GAD = Generalised Anxiety Disorder; ADHD = Attention Deficit Hyperactivity
Disorder; ODD = Oppositional Defiant Disorder.

### Associations between stressful life events at 7-8 years and wetting at 9 years

We found weak evidence of a sex interaction with stressful life events (OR (95% CI) 1.89 (0.96, 3.70), p=0.065). Consequently, we report the results for males and females separately (Table 3). Across all levels of adjustment there was evidence for an association between stressful life events in females at 7-8 years and wetting at 9 years (fully adjusted model OR (95% CI) = 1.66 (1.05, 2.61), p=0.029), but no evidence for an association in males (fully adjusted model OR (95% CI) = 0.87 (0.52, 1.47), p=0.608). There was little evidence for a sex interaction in complete case results (Table S6).

**Table 3.** Associations between stressful life events aged 7-8 years and relapse of wetting at 9 years (imputed data, n=6408)

|  | Adjusted for sex |  | Adjusted for sex and socioeconomic disadvantage |  | Adjusted for sex, socioeconomic disadvantage, and maternal mental health |  | Adjusted for sex, socioeconomic disadvantage, maternal mental health, child developmental level, and child mental health |  |
| --- | --- | --- | --- | --- | --- | --- | --- | --- |
|  | OR (95%CI) | p | OR (95%CI) | p | OR (95%CI) | p | OR (95%CI) | p |
| **Male** | 0.96 (0.58, 1.59) | .864 | 0.94 (0.56, 1.57) | .818 | 0.90 (0.54, 1.52) | .701 | 0.87 (0.52, 1.47) | .608 |
| **Female** | 1.81 (1.16, 2.81) | .009 | 1.71 (1.09, 2.69) | .019 | 1.67 (1.07, 2.62) | .025 | 1.66 (1.05, 2.61) | .029 |
| **Sex interaction** | 1.89 (0.96, 3.70) | .065 | 1.82 (0.93, 3.58) | .082 | 1.85 (0.94, 3.65) | .076 | 1.90 (0.96, 3.76) | .066 |

## Discussion

This is the largest study to date analysing the prospective association of symptoms of common mental health disorders (7 years) and stressful life events (7-8 years) with subsequent relapse in UI (9 years). The presence of separation anxiety symptoms at age 7 was strongly associated with a relapse in UI, and this association remained in the fully adjusted models. There was some evidence that social anxiety, ADHD and ODD symptoms were associated with relapse in UI, but these associations attenuated following adjustment for child developmental level and earlier emotional/behaviour problems. There was weak evidence for an interaction between stressful life events and sex; females who experienced three or more stressful life events had increased odds of relapse, but there was no association in males. This pattern was consistent across all levels of adjustment for confounders.

### Strengths and limitations

To our knowledge this is the largest study examining the prospective associations between symptoms of mental health disorders, stressful life events and relapse in UI in children. Additional strengths include the examination of a range of common mental disorder symptoms and stressful life events preceding relapse in UI, adjustment for a comprehensive list of potential confounders, and use of multiple imputation to address possible bias due to missing data.

We used maternal reports of symptoms of mental health disorders (any severe symptoms) and UI (any wetting) and did not restrict our analysis to children who met clinical diagnostic criteria. The study, therefore, provides evidence that symptoms of mental health disorders are associated with relapse in UI in a non-clinical sample of children in the community.

We were unable to examine whether symptoms of mental health disorders and stressful life events were more strongly associated with a relapse in bedwetting or daytime wetting due to small numbers of children who experienced relapse in different subtypes of wetting at age 9.

We did not examine specific stressful life events. It is possible that particular stressful life events may be more, or less, associated with subsequent wetting [15]. Different types of stressful life events, however, are likely to be interrelated (e.g. being taken into care, separation from mother/father or a change in caregiver) and specific types of stressful events show moderate correlations in ALSPAC, for instance sexual and physical abuse [23]. Previous research examining single stressful life events (e.g. parental divorce and separation) in ALSPAC found no evidence for individual associations with bedwetting [6]. We chose to dichotomise a total score rather than focus on specific events as evidence suggests that the total burden of stressful life events is an important risk factor for wetting rather than any specific event in isolation [6, 24].

Finally, the ALSPAC sample is predominantly white and affluent and under-representative of the population at the time [16, 17]. We are therefore unable to generalise our results to minority ethnic groups and less affluent populations. Further research in these under-served populations is vital to prevent widening inequalities in heath research.

### Comparison with previous studies

When all confounders are taken into consideration, separation anxiety symptoms remained the only mental health symptoms at age 7 that were associated with a relapse in UI at age 9. This underlines the importance of anxiety disorders in the aetiology of wetting, which have been neglected in previous research (Hussong et al., under review). In the study of Hussong et al. (under review) separation anxiety was the most common anxiety disorder among children with incontinence.

Separation anxiety carries a genetic disposition and can be induced by stressful life events [25]. It could be speculated that the same environmental stressors could increase the risk for separation anxiety, as well as for relapses in UI.

Although our confidence intervals covered a potential detrimental effect of behavioural problems (ODD, ADHD, conduct disorder symptoms) on relapse of UI, our point estimates indicated there was no evidence of strong associations. This is different to McGee et al. [13] and Feehan et al. [14] who found higher levels of behavioural problems in secondary enuresis compared to no enuresis.

Importantly, we adjusted for a range of confounders including child developmental level and early mental health problems which may explain these differences. Our results are consistent with Feehan et al.’s finding of higher anxiety scores in children with secondary enuresis compared with dry children [14].

We found that exposure to three or more stressful life events was associated with subsequent relapse in females only. Previous studies found that stressful life events were associated with secondary enuresis in both sexes [8, 15]. The inconsistent results might be due to different definitions of relapse between studies. Jarvelin et al. focused on relapse in day or night wetting at age 7 after at least 6 months of dryness, and Fergusson et al. examined relapse in night wetting at 10 years after at least 1 year of dryness. Importantly, our study was the first to examine sex differences in the association between stressful life events and relapse in UI. There is evidence for sex differences in sensitivity to stressful life events, as girls are more susceptible to stress and more likely to experience stressful events than boys [26–28]. Our results suggest that relapse in UI may be one outcome of this sensitivity to stress in females. Further studies are needed to substantiate the evidence for this sex difference.

### Possible mechanisms explaining the findings

Enuresis is understood as a genetically determined developmental disorder, which is modulated by environmental factors; daytime wetting is also multifactorial with a complex interaction of genetic and environmental factors [9, 29–31]. There is evidence for genetic overlap of enuresis with ADHD and affective disorders [33]. However, it is unclear whether this is due to the same set of risks manifesting as different disorders in different people, at different ages, or because ADHD/affective disorders causes enuresis (or both). Distinguishing between these explanations, using causal inference methods such as Mendelian randomization analysis [34], is important because there are different prevention and treatment implications.

It is possible that shared genetic and environmental factors for mental disorders also contribute to relapse in UI. Maternal anxiety is believed to be associated with child anxiety through mechanisms including genetic transmission, biological effects of stress in utero, and parenting factors such as critical or overcontrolling parenting styles [35]. It is plausible that these factors lead to child stress which, in turn, could lead to child anxiety and/or UI. Evidence suggests that maternal anxiety is also associated with subsequent child UI [36, 37] but there has been limited research on the mechanisms through which this association occurs. Preliminary evidence suggests that the mechanism is likely to occur in the postnatal rather than the antenatal period. Sawyer et al. (under review) found evidence for an association between maternal postnatal anxiety and offspring incontinence at age 7 but no evidence for a causal inter-uterine effect using a negative control design.

Presence of separation anxiety symptoms and stressful life events may result in an accumulation of psychological stress which impacts the bladder. Animal studies have found that psychological stress can increase voiding frequency and bladder dysfunction [38]. Dysregulation of the hypothalamic-pituitary-adrenal (HPA) axis is one potential mechanism, with animal studies indicating evidence for functional alterations such as increased corticotropin-releasing factor (CRF) [39, 40], and human studies showing alterations in brain activation in regions such as the anterior cingulate gyrus for women experiencing increased urinary urgency [41, 42]. Other potential mechanisms include increased mast cells and angiogenesis (formation of new blood vessels) [43], alterations in neurotransmitters such as serotonin, dopamine and glutamate [44–46], and bladder hypersensitivity [38]. Chronic stress has different biological effects on micturition in male and female rats, with differences in oestrogen and the HPA-axis being hypothesised as a key mechanisms [38]. These may be underlying factors for sex differences in the association between stressful life events and relapse in UI we observed in the current study.

## Conclusions

Our findings suggest that intervening early in childhood to treat separation anxiety and to reduce the negative outcomes associated with stressful life events may help to reduce the risk of relapse in UI. Replication in independent samples and studies using methods to improve causal inference are required to substantiate these findings. Clinicians should also be aware that girls experiencing stressful life events may be at greater risk for relapse in UI. Although stressful life events are hard to prevent, it might be possible to intervene on modifiable factors on the causal pathway. Further research is required to examine whether these associations are causal, and to identify potential underlying mechanisms that could be targeted in interventions.

## Data Availability

ALSPAC data access is through a system of managed open access. The steps below highlight how to apply for access to the data included in this paper and all other ALSPAC data.
1. Please read the ALSPAC access policy (http://www.bristol.ac.uk/media-library/sites/alspac/documents/researchers/data-access/ALSPAC_Access_Policy.pdf) which describes the process of accessing the data and samples in detail, and outlines the costs associated with doing so.
2. You may also find it useful to browse our fully searchable research proposals database, which lists all research projects that have been approved since April 2011.
3. Please submit your research proposal (https://proposals.epi.bristol.ac.uk/) for consideration by the ALSPAC Executive Committee. You will receive a response within 10 working days to advise you whether your proposal has been approved.
If you have any questions about accessing data or samples, please (data) or bbl-info{at}bristol.ac.uk (samples).

## Acknowledgments

We are extremely grateful to all the families who took part in this study, the midwives for their help in recruiting them, and the whole Avon Longitudinal Study of Parents and Children team, which includes interviewers, computer and laboratory technicians, clerical workers, research scientists, volunteers, managers, receptionists and nurses.

## Conflicts of interest

The authors have no conflicts of interest to declare that are relevant to the content of this article.

## Author Contributions Statement

Conceptualization: Carol Joinson, Jon Heron, Naomi Warne; Formal analysis: Naomi Warne, Jon Heron; Supervision: Carol Joinson, Jon Heron; Writing – original draft: Naomi Warne, Carol Joinson; Writing – review & editing: all authors.

## Availability of data

ALSPAC data access is through a system of managed open access. The steps below highlight how to apply for access to the data included in this paper and all other ALSPAC data.

1. Please read the <u>ALSPAC access policy (PDF, 891kB)</u> which describes the process of accessing the data and samples in detail, and outlines the costs associated with doing so.
2. You may also find it useful to browse our fully searchable <u>research proposals database</u>, which lists all research projects that have been approved since April 2011.
3. Please <u>submit your research proposal</u> for consideration by the ALSPAC Executive Committee. You will receive a response within 10 working days to advise you whether your proposal has been approved.

If you have any questions about accessing data or samples, please (data) or (samples).

## Notes

Funding This work is supported by funding from the Medical Research Council (grant ref: MR/V033581/1: Mental Health and Incontinence). The UK Medical Research Council and Wellcome (Grant ref: 217065/Z/19/Z) and the University of Bristol provide core support for ALSPAC. This publication is the work of the authors and Naomi Warne, Jon Heron and Carol Joinson will serve as guarantors for the contents of this paper. A comprehensive list of grants funding is available on the ALSPAC website (http://www.bristol.ac.uk/alspac/external/documents/grant-acknowledgements.pdf).

### Competing Interest Statement

The authors have declared no competing interest.

### Funding Statement

This work is supported by funding from the Medical Research Council (grant ref: MR/V033581/1: Mental Health and Incontinence). The UK Medical Research Council and Wellcome (Grant ref: 217065/Z/19/Z) and the University of Bristol provide core support for ALSPAC. This publication is the work of the authors and Naomi Warne, Jon Heron and Carol Joinson will serve as guarantors for the contents of this paper. A comprehensive list of grants funding is available on the ALSPAC website (http://www.bristol.ac.uk/alspac/external/documents/grant-acknowledgements.pdf).

### Author Declarations

The Avon Longitudinal Study of Parents and Children Ethics and Law Committee of University of Bristol and the Local Research Ethics Committees gave ethical approval for this work.

