## Supplementary material for "Mental health problems, stressful life events and relapse in urinary incontinence in primary school-age childhood: A prospective cohort study": Table S

Supplementary Table 1. Differences in confounders for those included in the sample (dry at 7 years) and those excluded from the sample (wet at 7 years)

| Confounder | Value | | Wet at 7 | Dry at 7 | Test statistic | p value |
|---|---|---|---|---|---|---|
| Sex (n=8018) | 0 (male) | n | 967 | 3,143 | 62.48 | <.001 |
| | | % | 60.06 | 49.05 | | |
| | 1 (female) | n | 643 | 3,265 | | |
| | | % | 39.94 | 50.95 | | |
| Social class (n=7416) | 0 (non-manual) | n | 1258 | 5053 | 0.37 | 0.545 |
| | | % | 84.60 | 85.23 | | |
| | 1 (manual) | n | 229 | 876 | | |
| | | % | 15.4 | 14.77 | | |
| Maternal age (n=8018) | 0 (20 or older) | n | 1574 | 6264 | <0.01 | 0.978 |
| | | % | 97.76 | 97.75 | | |
| | 1 (19 or younger) | n | 36 | 144 | | |
| | | % | 2.24 | 2.25 | | |
| Parity (n=7775) | 0 (0-2) | n | 1,481 | 5,925 | 1.08 | 0.298 |
| | | % | 94.75 | 95.38 | | |
| | 1 (3+) | n | 82 | 287 | | |
| | | % | 5.25 | 4.62 | | |
| Maternal education (n=7785) | 0 (A level or greater) | n | 700 | 2,530 | 7.95 | 0.019 |
| | | % | 44.61 | 40.70 | | |
| | 1 (O level) | n | 520 | 2,221 | | |
| | | % | 33.14 | 35.73 | | |
| | 2 (vocational/less) | n | 349 | 1,465 | | |
| | | % | 22.24 | 23.57 | | |
| Home ownership status (n=7172) | 0 (Mortgage/ owned/ private rented) | n | 1259 | 5038 | 0.06 | 0.809 |
| | | % | 87.61 | 87.85 | | |
| | 1 (Rent/ other) | n | 178 | 697 | | |
| | | % | 12.39 | 12.15 | | |
| Major financial difficulties (n=7120) | 0 | n | 1270 | 5191 | 17.60 | <.001 |
| | | % | 87.89 | 91.47 | | |
| | 1 | n | 175 | 484 | | |
| | | % | 12.11 | 8.53 | | |
| Material hardship (n=7044) | range 0-15 | n | 1406 | 5638 | 3.96 | 0.0001 |
| | | mean | 2.37 | 2.03 | | |
| | | SD | 3.05 | 2.84 | | |
| Social support (n=7089) | Range 1-30 | n | 1436 | 5,653 | 3.30 | 0.001 |
| | | mean | 19.81 | 20.29 | | |
| | | SD | 5.07 | 5.00 | | |
| Maternal depression (n=7100) | Range 0-29 | n | 1440 | 5660 | 4.93 | <.001 |
| | | mean | 6.83 | 6.09 | | |
| | | SD | 5.25 | 5.06 | | |
| Maternal anxiety (n=7066) | Range 0-16 | n | 1430 | 5636 | 6.30 | <.001 |
| | | mean | 5.62 | 4.95 | | |
| | | SD | 3.71 | 3.55 | | |

| | | | | | | |
|---|---|---|---|---|---|---|
| Child developmental level (n=6122) | Range -1.29 - 3.32 | n | 1229 | 4893 | 6.72 | <.001 |
| | | mean | -0.16 | 0.04 | | |
| | | SD | 1.01 | 0.94 | | |
| Child mental health (SDQ) (n=7131) | Range 0-31 | n | 1427 | 5704 | 11.56 | <.001 |
| | | mean | 8.75 | 7.12 | | |
| | | SD | 5.09 | 4.67 | | |
| Child stressful life events (n=7196) | Range 0-17 | n | 1441 | 5755 | 4.65 | <.001 |
| | | mean | 1.51 | 1.32 | | |
| | | SD | 1.54 | 1.37 | | |

SDQ = Strengths and Difficulties Questionnaire

Supplementary Table 2. Variable derivation for mental health exposures on the Development and Wellbeing Assessment (Goodman, Ford, Richards, Gatward, & Meltzer, 2000)

| Variable name | Symptoms | Frequency/Response | Final variable coding |
|---|---|---|---|
| ***Separation anxiety*** | | | |
| KR201 | Child worried about something unpleasant happening to special people in past month relative to peers | No more than others<br>A little more than others<br>A lot more than others | If any symptom was rated "a lot more than others" then separation anxiety symptom = 1 (yes) |
| KR202 | Child worried about being taken away from special people in past month relative to peers | | |
| KR203 | Child was unwilling to go to school in case something nasty happened to special people in past month relative to peers | | |
| KR204 | Child worried about sleeping alone in past month relative to peers | | |
| KR205 | Child left bedroom at night to check on special people in past month relative to peers | | |
| KR206 | Child worried about sleeping in a strange place in past month relative to peers | | |
| KR207 | Child was afraid of being alone in a room at home without special people in past month relative to peers | | |
| KR208 | Child had repeated nightmares about being separated from special people in past month relative to peers | | |
| KR209 | Child was ill when leaving special people in past month relative to peers | | |
| KR210 | Child had tantrums when separated from special people in past month relative to peers | | |
| ***Phobias*** | | | |
| KR225 | Child is scared of insects, spiders, wasps, bees, mice, snakes, birds or any other creature | Not at all<br>Only a little<br>Quite a lot<br>A great deal | If any symptom was rated "a great deal" then phobia = 1 (yes) |
| KR226 | Child is scared of storms, thunder, water or heights | | |
| KR227 | Child is scared of blood, injections or injury | | |
| KR228 | Child is scared of doctors or dentists | | |
| KR229 | Child is scared of other specific situations: lifts, tunnels, flying, driving, trains buses, small enclosed spaces | | |
| KR230 | Child is scared of the dark | | |
| KR231 | Any other specific fear | | |
| ***Social anxiety*** | | | |
| KR251 | Child was afraid of meeting new people in past month | Not at all | |

| Variable name | Symptoms | Frequency/Response | Final variable coding |
|---|---|---|---|
| KR252 | Child was afraid of meeting lots of people in past month | A little<br>A lot<br>Not done this | If any symptom was rated "a lot" then social anxiety = 1 (yes) |
| KR253 | Child was afraid of speaking in class in past month | | |
| KR254 | Child was afraid of reading out loud before others in past month | | |
| KR255 | Child was afraid of writing in front of others in past month | | |
| KR256 | Child was afraid of eating in front of others in past month | | |
| ***Post-Traumatic Stress Disorder*** | | | |
| KR285 | Child relived stressful event with vivid memories in past month | Not at all<br>A little<br>A lot | If any symptom was rated "a lot" then PTSD = 1 (yes)<br>Note: These questions were asked following a screener question *"During your study child's lifetime has anything exceptionally stressful happened to her, that would really upset almost anyone, such as being involved in a terrible accident, or being abused or some other sort of disaster?"*<br>If no to screener, then PTSD = 0 |
| KR286 | Child had repeated bad dreams of stressful event in past month | | |
| KR287 | Child got upset by reminders of stressful event in past month | | |
| KR288 | Child avoided talking about stressful event in past month | | |
| KR289 | Child avoided activities/places/people related to stressful event in past month | | |
| KR290 | Child blocked out details of stressful event from memory in past month | | |
| KR291 | Child had reduced interest in activities in past month | | |
| KR292 | Child expressed reduced range of feelings in past month | | |
| KR293 | Child had problems sleeping in past month | | |
| KR294 | Child seemed irritable/angry in past month | | |
| KR295 | Child had difficulty concentrating in past month | | |
| KR296 | Child was always alert for possible dangers in past month | | |
| KR297 | Child was easily startled in past month | | |
| ***Obsessive Compulsive Disorder*** | | | |
| KR321 | Child has repeatedly cleaned self excessively in past month | Never<br>Sometimes<br>Often | If any symptom was rated "often" then OCD = 1 (yes) |
| KR322 | Child has repeatedly taken other special measures to avoid dirt in past month | | |
| KR323 | Child has repeatedly checked things in past month | | |
| KR324 | Child has repeatedly performed repeated actions in past month | | |
| KR325 | Child has repeatedly touched things/people in particular ways in past month | | |
| KR326 | Child has repeatedly arranged things symmetrically in past month | | |
| KR327 | Child has repeatedly counted to lucky numbers / avoided unlucky numbers in past month | | |
| ***Generalised Anxiety Disorder*** | | | |
| KR357 | Child worries about past behaviour | Never<br>Sometimes<br>Often | If any symptom was rated "often" then GAD = 1 (yes) |
| KR358 | Child worries about schoolwork/homework | | |
| KR359 | Child worries about disasters | | |
| KR360 | Child worries about health | | |

| Variable name | Symptoms | Frequency/Response | Final variable coding |
|---|---|---|---|
| KR361 | Child worries about bad things happening to others | | |
| KR362 | Child worries about the future | | |
| KR363 | Child worries about other things | | |
| ***Depression*** | | | |
| KR410 | Child lacked energy in past month | Yes<br>No | |
| KR411 | Child ate much more/less than usual in past month | | If any symptom was rated "yes" then depression = 1 (yes)<br>Note: These questions were asked following three screener questions:<br>*In the past month, have there been times when your study child:*<br>*1) has been very sad, miserable, unhappy or tearful?*<br>*2) has been grumpy or irritable in a way that was out of character for her?*<br>*3) lost interest in everything, or nearly everything, she normally enjoys doing?*<br>If no to screeners, then depression = 0 |
| KR412 | Child lost/gained a lot of weight in past month | | |
| KR413 | Child found it hard to get to sleep in past month | | |
| KR414 | Child slept too much in past month | | |
| KR415 | Child was frequently agitated for a period in past month | | |
| KR416 | Child frequently felt worthless/guilty for a period in past month | | |
| KR417 | Child found it unusually hard to concentrate for a period in past month | | |
| KR418 | Child thought about death a lot in past month | | |
| KR419 | Child talked about harming/killing self in past month | | |
| KR420 | Child tried to harm/kill self in past month | | |
| ***Attention Deficit Hyperactivity Disorder*** | | | |
| KR436 | Child often fidgeted in past 6 months relative to peers | No more than others<br>A little more than others<br>A lot more than others | If any symptom was rated "a lot more than others" then ADHD = 1 (yes) |
| KR437 | Child found it hard to sit down for long in past 6 months relative to peers | | |
| KR438 | Child ran or climbed about illicitly in past 6 months relative to peers | | |
| KR439 | Child found it hard to play quietly in past 6 months relative to peers | | |
| KR440 | Child found it hard to calm down in past 6 months relative to peers | | |
| KR441 | Child often blurted out answers in past 6 months relative to peers | | |
| KR442 | Child found it hard to wait own turn in past 6 months relative to peers | | |
| KR443 | Child often butted into conversations/games in past 6 months relative to peers | | |
| KR444 | Child often went on talking when asked to stop in past 6 months relative to peers | | |
| KR448 | Child often made careless mistakes in past 6 months relative to peers | | |
| KR449 | Child often lost interest in activities in past 6 months relative to peers | | |
| KR450 | Child often didn't listen when addressed in past 6 months relative to peers | | |
| KR451 | Child often didn't finish a job properly in past 6 months relative to peers | | |
| KR452 | Child often found it hard to get organised in past 6 months relative to peers | | |

| Variable name | Symptoms | Frequency/Response | Final variable coding |
|---|---|---|---|
| KR453 | Child often tried to get out of activities involving thought in past 6 months relative to peers | | |
| KR454 | Child often lost things needed for school in past 6 months relative to peers | | |
| KR455 | Child was easily distracted in past 6 months relative to peers | | |
| KR456 | Child was often forgetful in past 6 months relative to peers | | |
| ***Oppositional Defiant Disorder*** | | | |
| KR481 | Child has had severe temper tantrums in past 6 months relative to peers | No more than others<br>A little more than others<br>A lot more than others | If any symptom was rated "a lot more than others" then ODD = 1 (yes) |
| KR482 | Child has argued with grown-ups in past 6 months relative to peers | | |
| KR483 | Child has ignored rules / been disobedient in past 6 months relative to peers | | |
| KR484 | Child has deliberately annoyed people in past 6 months relative to peers | | |
| KR485 | Child has blamed others for own mistakes in past 6 months relative to peers | | |
| KR486 | Child has been easily annoyed in past 6 months relative to peers | | |
| KR487 | Child has been angry & resentful in past 6 months relative to peers | | |
| KR488 | Child has been spiteful in past 6 months relative to peers | | |
| KR489 | Child has tried to get revenge on people in past 6 months relative to peers | | |
| ***Conduct disorder*** | | | |
| KR503 | Child has told lies to obtain objects/favours or to avoid duties in past 12 months | No<br>Perhaps<br>Definitely | If any symptom was rated "definitely" or "More than once" then conduct = 1 (yes) |
| KR505 | Child has started fights with non-siblings in past 12 months | | |
| KR507 | Child has bullied/threatened people in past 12 months | | |
| KR509 | Child has stayed out later than allowed in past 12 months | | |
| KR511 | Child has stolen things in past 12 months | | |
| KR515 | Child has often played truant in past 12 months | | |
| KR513 | Child has run away / stayed away all night without permission in past 12 months | Never<br>Once only<br>More than once | |

Supplementary material for: *Mental health problems, stressful life events and relapse in urinary incontinence in primary school-age childhood: A prospective cohort study*

Supplementary Table 3. Variable creation for confounders

| **Measure/questionnaire** | **Variable name** | **Question** | **Responses** | **Final variable coding** |
|---|---|---|---|---|
| ***Child sex*** | | | | |
| Recorded at birth by the fieldworkers who visited the maternity units | kz021 | n/a | 1- male<br>2 – female<br>-1 - not known | 0 male<br>1 female<br>(not known = missing) |
| ***Social class (sociodemographic indicator)*** | | | | |
| 1991 British Office of Population and Census Statistics (OPCS) job codes (maternal) | c755 | Derived variable from questions:<br>• Actual job, occupation, trade or profession<br>• Please tick which of the following apply to you: foreman, manager, supervisor, leading hand, self-employed, none of these<br>• Type of industry or service given (main things done in job): | 1 - I<br>2 - II<br>3 – III (non-manual)<br>4 – III (manual)<br>5 - IV<br>6 - V<br>65 - Armed forces<br>-1 - missing | Highest parental social class *i.e. if one parent is non-manual then social class=0*:<br>0 = non-manual: professional, managerial or skilled professions (responses 1, 2, 3)<br>1 = manual: partly or unskilled occupations (responses 4, 5, 6)<br>65 = missing<br>-1 = missing |
| 1991 British Office of Population and Census Statistics (OPCS) job codes (paternal) | c765 | | | |
| ***Mother's age at birth (sociodemographic indicator)*** | | | | |
| | mz028b | Derived variable: Grouped age of mother at delivery | Continuous variable <16 through to >43 (-10, -4, -2 missing) | 0 = 20 or older<br>1 = 19 or younger |
| ***Parity (sociodemographic indicator)*** | | | | |
| | b032 | Derived variable: number of previous pregnancies resulting in either a livebirth or a stillbirth | Continuous score (range 0-22) (-7, -2, -1 missing) | 0 = less than 3 children<br>1 = 3 or more children |
| ***Maternal education (sociodemographic indicator)*** | | | | |
| | c645a | Derived variable: mum's highest educational qualification | 1 – CSE/none<br>2 – Vocational<br>3 – O level<br>4 – A level<br>5 – degree<br>-1 – missing | 0 = A level or greater<br>1 = O level<br>2 = vocational or less |
| ***Home ownership (sociodemographic indicator)*** | | | | |
| | k5010 | Is your home:<br>a) being bought/mortgaged | 0 Being bought/mortgaged<br>1 Being bought from council | 0 = Mortgage/ owned/ privately rented (responses 0, 1, 2, 4, 5) |

| Measure/questionnaire | Variable name | Question | Responses | Final variable coding |
|---|---|---|---|---|
| | | b) being bought from council<br>c) owned - with no mortgage to pay<br>d) rented from council<br>e) rented from private landlord - furnished<br>f) rented from private landlord - unfurnished<br>g) rented from housing association<br>h) other (please tick & describe) | 2 Owned with no mortgage to pay<br>3 Rented from council<br>4 Rented from private landlord furnished<br>5 Rented from private landlord unfurnished<br>6 Rented from housing association<br>7 other | 1 = rent/other (responses 3, 6, 7) |
| ***Major financial difficulties (sociodemographic indicator)*** | | | | |
| ALSPAC life events inventory questionnaire previously used in (Joinson, Kounali, & Lewis, 2017) | I4024 | Listed below are a number of events which may have brought changes in your life. Have any of the these occurred since your study child's 5th birthday?<br>Since your child was 5:<br>You had a major financial problem | 1 Yes & affected respondent a lot<br>2 Yes, moderately affected<br>3 Yes, mildly affected<br>4 Yes, did not affect respondent at all<br>5 No, did not happen | 0 = no (response 5)<br>1 = yes (responses 1 through 4) |
| ***Material hardship (sociodemographic indicator)*** | | | | |
| ALSPAC hardship items previously used in (Joinson et al., 2017) | <br><br>K6200<br>K6201<br>K6202<br>K6203<br>K6204 | How difficult at the moment do you find it to afford these items:<br>a) food<br>b) clothing<br>c) heating*<br>d) rent or mortgage*<br>e) things you need for your children | 1 Very difficult<br>2 Fairly difficult<br>3 Slightly difficult<br>4 Not difficult<br>5 Paid directly by social security | Recode K6200-K6204 (5=4)<br>20 - K6200 - K6201 - K6202 - K6203 - K6204<br>Creates a summed score 0-15 (higher scores = more material hardship) |
| ***Social support (sociodemographic indicator)*** | | | | |
| ALSPAC social support scale previously used in (Tracy, Salo, & Appleton, 2018) | L7020 | I have no one to share my feelings with | 1 – Exactly feel<br>2 – Often feel<br>3 - Sometimes feel<br>4 - Never feel<br>*7 – No partner (response only for items specifically about partner) | Summed score with reverse coding for some items<br>Recode:<br>L7020: (1=0)(2=1)(3=2)(4=3)<br>L7021: (1=3)(2=2)(3=1)(4=0)(7=3)<br>L7024: (1=0)(2=1)(3=2)(4=3) (7=3)<br>L7026: (1=3)(2=2)(3=1)(4=0)(7=3) |
| | L7021 | My partner provides the emotional support I need* | | |
| | L7022 | There are other mothers with whom I can share my experiences | | |
| | L7023 | I believe in moments of difficulty my neighbours would help me | | |
| | L7024 | I'm worried that my partner might leave me* | | |

| Measure/questionnaire | Variable name | Question | Responses | Final variable coding |
|---|---|---|---|---|
| | L7025 | There is always someone with whom I can share my happiness and excitement about my child | | L7022- L7023, L7025, L2027- L7029: (1=3)(2=2)(3=1)(4=0)<br>Sum all recoded items (higher scores = more social support) |
| | L7026 | If I feel tired I can rely on my partner to take over* | | |
| | L7027 | If I was in financial difficulty I know my family would help if they could | | |
| | L7028 | If I was in financial difficulty I know my friends would help if they could | | |
| | L7029 | If all else fails I know the state will support and assist me | | |
| ***Maternal depression*** | | | | |
| Edinburgh Postnatal Depression Scale(Cox, Holden, & Sagovsky, 1987) | | Your feelings in the past week: / In the past week: | | |
| | l2010 | I have been able to laugh and see the funny side of things: | 1 – As much as I always could<br>2 – Not quite so much now<br>3 – Definitely not so much now<br>4 - Not at all | Recode l2010, l2011, l2013 (1=0)(2=1)(3=2)(4=3)<br>Recode l2012, l2014-l2019 (1=3)(2=2)(3=1)(4=0)<br>Any missing values then set new variable to missing<br>Sum all 10 items |
| | l2011 | I have looked forward with enjoyment to things | 1 - As much as I ever did<br>2 – Rather less than I used to<br>3 – Definitely less than I used to<br>4 - Hardly at all | |
| | l2012 | I have blamed myself unnecessarily when things went wrong | 1 – Yes, most of the time<br>2 – Yes, some of the time<br>3 – Not very often<br>4 - Never | |
| | l2013 | I have been anxious or worried for no good reason | 1 – No, not at all<br>2 – Hardly ever<br>3 – Yes, sometimes<br>4 – Yes, often | |
| | l2014 | I have felt scared or panicky for no good reason | 1 – Yes, quite a lot<br>2 – Yes, sometimes<br>3 – No, not much<br>4 – No, not at all | |
| | l2015 | Things have been getting on top of me | 1 – Yes, most of the time I haven't been able to cope | |

| Measure/questionnaire | Variable name | Question | Responses | Final variable coding |
|---|---|---|---|---|
| | | | 2 – Yes, sometimes I haven't been coping as well as usual<br>3 – No, most of the time I have coped quite well<br>4 - No, I have been coping as well as ever | |
| | l2016 | I have been so unhappy that I have had difficulty sleeping | 1 – Yes, most of the time<br>2 – Yes, sometimes<br>3 – Not very often<br>4 - No, not at all | |
| | l2017 | I have felt sad or miserable | 1 – Yes, most of the time<br>2 – Yes, sometimes<br>3 – Not very often<br>4 - No, not at all | |
| | l2018 | I have been so unhappy that I have been crying | 1 – Yes, most of the time<br>2 – Yes, quite often<br>3 – Only occasionally<br>4 - Never | |
| | l2019 | The thought of harming myself has occurred to me | 1 – Yes, quite often<br>2 – Sometimes<br>3 – Hardly ever<br>4 – Never | |
| ***Maternal anxiety*** | | | | |
| Crown Crisp Experimental Index anxiety subscale (Crown & Crisp, 1979) | L2000 | Do you feel upset for no obvious reason? | 1 – Very often<br>2 – Often<br>3 – Not very often<br>4 - Never | Recode l2000, l2005, l2006 1/2=2, 3/4=0<br>Recode l2003 1/3=2, 4=0<br>Recode l2001, l2002, l2004, l2007 1/2=2, 3=1, 4=0<br><br>Sum all 8 recoded items |
| | L2001 | Have you felt as though you might faint? | | |
| | L2002 | Do you feel uneasy and restless? | | |
| | L2003 | Do you sometimes feel panicky? | | |
| | L2004 | Do you worry a lot? | | |
| | L2005 | Do you feel strung-up inside? | | |
| | L2006 | Do you ever have the feeling you are going to pieces? | | |
| | L2007 | Do you have bad dreams which upset you when you wake up? | | |

| Measure/questionnaire | Variable name | Question | Responses | Final variable coding |
|---|---|---|---|---|
| ***Child developmental level*** | | | | |
| Adapted Denver Developmental Screening Test (Frankenburg & Dodds, 1967) | kd680 | Derived variable: Total ALSPAC development score: 18 months: Complete cases | Continuous score (range -7 to 2)<br>-100 = missing | Continuous score<br>(-100=missing) |
| ***Early child mental health*** | | | | |
| Strengths and Difficulties Questionnaire (Goodman, 2001) | kq348f | Derived variable: SDQ total difficulties score (prorated). | Continuous score (range 0-40) | Continuous score |
| ***Child stressful life events (confounder for DAWBA exposures only)*** | | | | |
| ALSPAC life events inventory questionnaire previously used in (Collin et al., 2015) | | Derived variables: since her 5th birthday | | |
| | kq360a | She was taken into care | 1 – Yes<br>2 – No<br><br>-8 – see text<br>-6 – Section D omitted<br>-1 - omitted | Recode (2=0)(1=1)(-8/-1 = missing)<br>Then sum score (possible range 0-17)<br>If half or more of items missing, then set to missing (in line with DAWBA exposure variables) |
| | kq361a | A pet died | | |
| | kq362a | She moved home | | |
| | kq363a | She had a shock or fright | | |
| | kq364a | She was physically hurt by someone | | |
| | kq365a | She was sexually abused | | |
| | kq366a | Somebody in the family died | | |
| | kq367a | She was separated from her mother | | |
| | kq368a | She was separated from her father | | |
| | kq369a | She acquired a new mother or father | | |
| | kq370a | She had a new brother or sister | | |
| | kq371a | She was admitted to hospital | | |
| | kq372a | She changed care taker (i.e. the person mostly looking after her) | | |
| | kq373a | She was separated from someone else that she was close to | | |
| | kq374a | She started a new school or kindergarten | | |
| | kq375a | She started school | | |
| | kq376a | She lost her best friend | | |

Supplementary Table 4. Descriptive information on mental health symptoms, stressful life events and wetting in imputed data and observed data

| | **Total** | | | | | **Males** | | | | **Females** | | | |
|---|---|---|---|---|---|---|---|---|---|---|---|---|---|
| | **Imputed data (n=6408)** | | **All available observed data** | | | **Imputed data (n=3143)** | | **All available observed data** | | **Imputed data (n=3265)** | | **All available observed data** | |
| | **%** | **SE** | **%** | **n** | **Missing n** | **%** | **SE** | **%** | **n** | **%** | **SE** | **%** | **n** |
| **Separation anxiety** | 6.59 | 0.31 | 6.56 | 411/6261 | 147 | 6.84 | 0.45 | 6.84 | 210/3070 | 6.34 | 0.43 | 6.30 | 201/3191 |
| **Phobia** | 11.56 | 0.40 | 11.55 | 738/6391 | 17 | 10.45 | 0.55 | 10.44 | 327/3133 | 12.62 | 0.58 | 12.62 | 411/3258 |
| **Social anxiety** | 5.13 | 0.28 | 5.12 | 325/6342 | 66 | 6.05 | 0.43 | 6.05 | 188/3108 | 4.25 | 0.36 | 4.24 | 137/3234 |
| **PTSD** | 2.99 | 0.21 | 2.97 | 189/6358 | 50 | 2.77 | 0.29 | 2.76 | 86/3118 | 3.20 | 0.31 | 3.18 | 103/3240 |
| **OCD** | 7.12 | 0.33 | 7.06 | 445/6302 | 106 | 7.90 | 0.49 | 7.87 | 244/3099 | 6.36 | 0.43 | 6.28 | 201/3203 |
| **GAD** | 8.03 | 0.34 | 8.00 | 508/6350 | 58 | 8.32 | 0.50 | 8.27 | 258/3120 | 7.74 | 0.47 | 7.74 | 250/3230 |
| **Depression** | 11.12 | 0.40 | 11.02 | 686/6225 | 183 | 10.21 | 0.55 | 10.11 | 309/3057 | 12.00 | 0.58 | 11.90 | 377/3168 |
| **ADHD** | 13.13 | 0.42 | 13.07 | 827/6329 | 79 | 16.54 | 0.67 | 16.51 | 512/3102 | 9.86 | 0.53 | 9.76 | 315/3227 |
| **ODD** | 5.24 | 0.28 | 5.25 | 331/6300 | 108 | 6.89 | 0.45 | 6.91 | 214/3097 | 3.66 | 0.33 | 3.65 | 117/3203 |
| **Conduct disorder** | 5.85 | 0.30 | 5.83 | 368/6315 | 93 | 6.49 | 0.44 | 6.47 | 201/3107 | 5.22 | 0.39 | 5.21 | 167/3208 |
| **Stressful life events** | 17.77 | 0.52 | 17.55 | 964/5493 | 915 | 17.12 | 0.74 | 16.97 | 453/2670 | 18.40 | 0.73 | 18.10 | 511/2823 |
| **Wetting at 9** | 4.53 | 0.29 | 4.35 | 234/5378 | 1030 | 4.92 | 0.43 | 4.78 | 124/2592 | 4.16 | 0.38 | 3.95 | 110/2786 |

PTSD = Post-Traumatic Stress Disorder; OCD = Obsessive Compulsive Disorder; GAD = Generalised Anxiety Disorder; ADHD = Attention Deficit Hyperactivity Disorder; ODD = Oppositional Defiant Disorder; SE = standard error

Supplementary Table 5. Full results at each stage of adjustment of association between mental health symptoms and relapse of wetting in imputed data (n=6408)

| | **Unadjusted** | | **Model A** | | **Model B** | | **Model C** | | **Model D** | | **Model E** | |
|---|---|---|---|---|---|---|---|---|---|---|---|---|
| **Exposure** | **OR (95%CI)** | **p** | **OR (95%CI)** | **p** | **OR (95%CI)** | **p** | **OR (95%CI)** | **p** | **OR (95%CI)** | **p** | **OR (95%CI)** | **p** |
| **Separation anxiety** | 2.63 (1.79, 3.85) | <.001 | 2.62 (1.78, 3.84) | <.001 | 2.51 (1.70, 3.71) | <.001 | 2.31 (1.55, 3.44) | <.001 | 2.09 (1.39, 3.14) | <.001 | 2.08 (1.39, 3.13) | <.001 |
| **Phobia** | 1.13 (0.77, 1.67) | .537 | 1.14 (0.77, 1.69) | .507 | 1.07 (0.72, 1.58) | .750 | 1.02 (0.68, 1.51) | .936 | 0.90 (0.60, 1.36) | .624 | 0.90 (0.60, 1.35) | .617 |
| **Social anxiety** | 1.96 (1.24, 3.11) | .004 | 1.93 (1.22, 3.06) | .005 | 1.83 (1.15, 2.92) | .011 | 1.69 (1.06, 2.72) | .029 | 1.47 (0.90, 2.38) | .121 | 1.47 (0.90, 2.38) | .121 |
| **PTSD** | 1.19 (0.56, 2.52) | .649 | 1.20 (0.57, 2.54) | .637 | 1.01 (0.47, 2.18) | .976 | 0.91 (0.42, 1.98) | .819 | 0.79 (0.36, 1.75) | .562 | 0.77 (0.35, 1.70) | .516 |
| **OCD** | 0.94 (0.56, 1.56) | .797 | 0.93 (0.56, 1.54) | .766 | 0.85 (0.51, 1.42) | .527 | 0.80 (0.48, 1.34) | .393 | 0.67 (0.39, 1.14) | .137 | 0.66 (0.39, 1.13) | .132 |
| **GAD** | 1.22 (0.78, 1.91) | .377 | 1.22 (0.78, 1.90) | .385 | 1.16 (0.74, 1.81) | .528 | 1.06 (0.67, 1.67) | .799 | 0.93 (0.58, 1.48) | .745 | 0.92 (0.58, 1.47) | .731 |
| **Depression** | 1.41 (0.98, 2.05) | .066 | 1.43 (0.99, 2.07) | .060 | 1.32 (0.90, 1.92) | .151 | 1.23 (0.84, 1.81) | .279 | 1.12 (0.76, 1.65) | .560 | 1.12 (0.76, 1.65) | .580 |
| **ADHD** | 1.65 (1.18, 2.30) | .003 | 1.62 (1.16, 2.26) | .005 | 1.49 (1.06, 2.10) | .022 | 1.40 (0.99, 1.98) | .061 | 1.05 (0.71, 1.55) | .794 | 1.05 (0.71, 1.54) | .822 |
| **ODD** | 2.21 (1.43, 3.42) | <.001 | 2.16 (1.40, 3.35) | .001 | 1.94 (1.24, 3.04) | .004 | 1.82 (1.15, 2.86) | .010 | 1.36 (0.83, 2.23) | .222 | 1.35 (0.82, 2.21) | .237 |
| **Conduct disorder** | 1.26 (0.75, 2.13) | .376 | 1.25 (0.74, 2.11) | .397 | 1.11 (0.65, 1.89) | .702 | 1.03 (0.60, 1.76) | .924 | 0.82 (0.47, 1.43) | .482 | 0.81 (0.47, 1.41) | .454 |

Model A: adjusted for sex. Model B: adjusted for sex and socioeconomic disadvantage. Model C: adjusted for sex, socioeconomic disadvantage and maternal mental health. Model D: adjusted for sex, socioeconomic disadvantage, maternal mental health and child development and child mental health. Model E: fully adjusted for sex, socioeconomic disadvantage, maternal mental health, child development and child mental health, and child stressful life events.

PTSD = Post-Traumatic Stress Disorder; OCD = Obsessive Compulsive Disorder; GAD = Generalised Anxiety Disorder; ADHD = Attention Deficit Hyperactivity Disorder; ODD = Oppositional Defiant Disorder

Supplementary material for: *Mental health problems, stressful life events and relapse in urinary incontinence in primary school-age childhood: A prospective cohort study*

Supplementary Table 6. Full results at each stage of adjustment of association between all exposures and relapse of wetting in complete cases (n=3047 for mental health exposures, n=3152 for stressful life events)

| | Unadjusted | | Model A | | Model B | | Model C | | Model D | | Model E | |
|---|---|---|---|---|---|---|---|---|---|---|---|---|
| **Exposure** | **OR (95%CI)** | **p** | **OR (95%CI)** | **p** | **OR (95%CI)** | **p** | **OR (95%CI)** | **p** | **OR (95%CI)** | **p** | **OR (95%CI)** | **p** |
| **Separation anxiety** | 2.24 (1.35, 3.71) | .002 | 2.24 (1.35, 3.71) | .002 | 2.13 (1.27, 3.56) | .004 | 2.00 (1.19, 3.36) | .009 | 1.90 (1.12, 3.23) | .018 | 1.90 (1.12, 3.23) | .018 |
| **Phobia** | 1.06 (0.64, 1.76) | .809 | 1.08 (0.65, 1.80) | .756 | 1.01 (0.61, 1.69) | .958 | 0.99 (0.59, 1.65) | .957 | 0.91 (0.54, 1.52) | .708 | 0.90 (0.54, 1.52) | .706 |
| **Social anxiety** | 1.89 (1.06, 3.35) | .030 | 1.84 (1.03, 3.27) | .038 | 1.65 (0.92, 2.95) | .094 | 1.54 (0.85, 2.77) | .154 | 1.41 (0.77, 2.58) | .261 | 1.41 (0.77, 2.58) | .261 |
| **PTSD** | 1.17 (0.47, 2.93) | .739 | 1.20 (0.48, 3.02) | .694 | 1.00 (0.39, 2.55) | .997 | 0.90 (0.35, 2.32) | .829 | 0.82 (0.31, 2.14) | .679 | 0.81 (0.31, 2.14) | .673 |
| **OCD** | 0.81 (0.39, 1.68) | .574 | 0.81 (0.39, 1.67) | .564 | 0.74 (0.35, 1.54) | .414 | 0.70 (0.33, 1.45) | .334 | 0.61 (0.29, 1.31) | .207 | 0.61 (0.29, 1.31) | .206 |
| **GAD** | 1.30 (0.76, 2.22) | .339 | 1.30 (0.76, 2.22) | .339 | 1.23 (0.71, 2.11) | .461 | 1.13 (0.65, 1.95) | .673 | 1.07 (0.61, 1.89) | .804 | 1.07 (0.61, 1.89) | .806 |
| **Depression** | 1.26 (0.78, 2.03) | .337 | 1.30 (0.81, 2.09) | .283 | 1.17 (0.72, 1.90) | .528 | 1.12 (0.69, 1.82) | .653 | 1.07 (0.65, 1.75) | .801 | 1.07 (0.65, 1.75) | .802 |
| **ADHD** | 1.81 (1.19, 2.75) | .006 | 1.73 (1.13, 2.65) | .011 | 1.58 (1.02, 2.44) | .041 | 1.47 (0.95, 2.30) | .086 | 1.25 (0.76, 2.04) | .38 | 1.25 (0.76, 2.04) | .382 |
| **ODD** | 1.97 (1.09, 3.57) | .025 | 1.90 (1.04, 3.45) | .036 | 1.74 (0.95, 3.21) | .075 | 1.63 (0.88, 3.02) | .124 | 1.32 (0.67, 2.58) | .419 | 1.32 (0.67, 2.59) | .422 |
| **Conduct disorder** | 0.70 (0.30, 1.61) | .399 | 0.69 (0.30, 1.58) | .375 | 0.61 (0.26, 1.42) | .250 | 0.56 (0.24, 1.32) | .186 | 0.46 (0.19, 1.09) | .078 | 0.45 (0.19, 1.09) | .077 |
| **Stressful life events[a]** | 1.25 (0.83, 1.89) | .281 | 1.26 (0.84, 1.91) | .264 | 1.23 (0.81, 1.86) | .336 | 1.18 (0.77, 1.79) | .446 | 1.17 (0.77, 1.78) | .461 | | |

Model A: adjusted for sex. Model B: adjusted for sex and socioeconomic disadvantage. Model C: adjusted for sex, socioeconomic disadvantage and maternal mental health. Model D: adjusted for sex, socioeconomic disadvantage, maternal mental health and child development and child mental health. Model E: fully adjusted for sex, socioeconomic disadvantage, maternal mental health, child development and child mental health, and child stressful life events.

PTSD = Post-Traumatic Stress Disorder; OCD = Obsessive Compulsive Disorder; GAD = Generalised Anxiety Disorder; ADHD = Attention Deficit Hyperactivity Disorder; ODD = Oppositional Defiant Disorder;

[a]There was little evidence of interaction with sex (OR (95% CI) = 1.65 (0.72, 3.79), p = .235) so results displayed are associations without sex interactions

Supplementary material for: *Mental health problems, stressful life events and relapse in urinary incontinence in primary school-age childhood: A prospective cohort study*
